# Limited Role of Antithrombin Deficiency in Nephrotic Syndrome-Associated Hypercoagulopathy

**DOI:** 10.1101/2022.06.24.22276257

**Authors:** Eman Abdelghani, Amanda P. Waller, Katelyn J. Wolfgang, Joseph R. Stanek, Samir V. Parikh, Brad H. Rovin, William E. Smoyer, the PNRC Investigators, the NEPTUNE Investigators, Bryce A. Kerlin

**Author notes:** See Addendum 1 for Pediatric Nephrology Research Consortium (PNRC) Investigators and Affiliations. See Addendum 2 for <u>Nep</u>hrotic Syndrome S<u>tu</u>dy <u>Ne</u>twork (NEPTUNE) Investigators and Affiliations. Amanda P. Waller and Eman Abdelghani contributed equally to this work. Corresponding Author: Bryce A. Kerlin, MD, Center for Clinical & Translational Research, The Abigail Wexner Research Institute at Nationwide Children’s, 700 Children’s Dr., W325, Columbus, OH 43205.

## Abstract

**Background:** Nephrotic syndrome is associated with an acquired hypercoagulopathy thought to drive its predisposition for venous thromboembolism. Previous studies have suggested that urinary antithrombin loss leading to acquired antithrombin deficiency is the primary mechanism underlying this hypercoagulopathy, but this hypothesis has not been directly tested. The objectives of this study were to test the influence of antithrombin levels on hypercoagulopathy in nephrotic syndrome patient samples and perform meta-analyses to evaluate the likelihood of antithrombin deficiency in nephrotic syndrome patients.

**Design:** Samples from 3 independent nephrotic syndrome cohorts were analyzed. Antithrombin antigen and activity assays were performed using ELISA and amidolytic assays, respectively. Plasma thrombin generation, albumin, and urine protein-to-creatinine ratios were determined using established methods. Meta-analyses were performed by combining these new data with previously published data.

**Results:** Antithrombin levels were not consistently related to either plasma albumin or proteinuria. Antithrombin was quantitatively related to hypercoagulopathy in adult nephrotic syndrome. Whereas antithrombin activity was inconsistently associated with hypercoagulopathy in childhood nephrotic syndrome. Notably, hypercoagulopathy did not differ between patients with normal antithrombin levels and those with levels below the threshold used to define clinical antithrombin deficiency (<70%). Moreover, *ex vivo* antithrombin supplementation did not significantly alter hypercoagulopathy in antithrombin deficient plasma samples. Moreover, the meta-analyses demonstrated that antithrombin deficiency is not a uniform feature of nephrotic syndrome but may be more common in children than adults.

**Conclusions:** These data suggest that antithrombin deficiency plays only a limited role in the mechanisms underlying the acquired hypercoagulopathy of nephrotic syndrome. Moreover, antithrombin deficiency is not present in all nephrotic syndrome patients and is more likely in children than adults despite the higher risk for venous thromboembolism in adults than children. Future studies should focus on identifying antithrombin-independent mechanisms that better explain the pathophysiology of hypercoagulopathy in nephrotic syndrome.

## INTRODUCTION

Nephrotic syndrome (NS) is an important risk factor for venous thromboembolism (VTE) which is a leading cause of worldwide mortality.^1-7^ VTE survivors may suffer substantial morbidity including recurrent VTE, post-thrombotic syndrome, or chronic thromboembolic pulmonary hypertension.^8, 9^ NS is associated with an acquired hypercoagulopathy that is thought to be a key pathophysiologic driver of VTE risk and historical data suggest that VTE complicates 25% of adult and 3% of childhood NS cases.^3, 10-13^

The mechanisms underlying NS-associated hypercoagulopathy include urinary loss of anticoagulant and profibrinolytic proteins, accumulation of procoagulant proteins, thrombocytosis, and platelet hyper-reactivity.^3, 14^ We and others have thus utilized global hemostasis assays, such as thrombin generation and thromboelastography, to better understand the summative effects of these complex derangements on coagulation activity and thus quantify NS-hypercoagulopathy.^10-13, 15^ Marked urinary antithrombin (AT) loss resulting in acquired AT deficiency has often been proposed as an important mechanism underlying NS-hypercoagulopathy.^3, 16-18^ Intuitively, this hypothesis is attractive because familial AT deficiency is the most severe heritable thrombophilia described to date.^19^ However, evidence directly assessing the contribution of AT deficiency to NS-hypercoagulopathy is lacking. Some studies have attributed VTE during NS to AT deficiency.^20-22^ Others have reported VTE despite normal AT levels.^23-25^ Additional reports demonstrated the absence of VTE despite AT deficiency.^25, 26^ Meanwhile, several studies from other fields have demonstrated a relationship between thrombin generation and AT deficiency.^27-29^ We thus reanalyzed our thrombin generation data from three independent NS cohorts to evaluate the influence of AT levels on NS-hypercoagulopathy.^10, 11^

Previous studies have shown conflicting data on the prevalence of AT deficiency during NS. While some studies observed significantly diminished AT levels in NS patients, others have demonstrated normal or even increased AT levels.^20, 23, 30-36^ Clinically, AT levels are quantified using both immunoassays to determine protein (antigen) levels and functional assays to determine its inhibitory activity. In rat NS models, we previously demonstrated that AT antigen was not correlated with proteinuria or thrombin generation.^11, 15^ Meanwhile, deficient AT activity was observed only in the setting of extreme proteinuria, suggesting that a qualitative (but not quantitative) AT deficiency may develop with severe proteinuria.^15^ This observation conflicts with the dogma that AT deficiency is driven by urinary AT loss leading to quantitative deficiency.^2, 3, 6, 16-18^ We thus performed meta-analyses incorporating available published data along with AT levels from our 3 cohorts to determine the prevalence of AT deficiency in NS patients. For this analysis we examined the overall effect of NS on AT levels and reported these observations in relationship to AT levels <70% (<0.7 IU/mL), a clinically relevant AT deficiency cut-off.^20, 37^

## MATERIALS AND METHODS

### Study Subjects

Complete descriptions, including human subject’s protections, of the <u>Nep</u>hrotic Syndrome S<u>tu</u>dy <u>Ne</u>twork (NEPTUNE), Pediatric Nephrology Research Consortium (PNRC), and Columbus cohorts have been published previously and are summarized in **Table 1**.^10, 11, 38^ Briefly, all three cohorts enrolled patients with incident NS from whom sodium citrate anticoagulated plasma was biobanked at -80°C. We have previously published NS-hypercoagulopathy studies using these plasma samples from all three cohorts.^10, 11^

**Table 1:** Key characteristics of the NEPTUNE, Columbus, and PNRC study designs.

| **Cohort** | **NEPTUNE<sup>10</sup>** | **PNRC<sup>11</sup>** | **Columbus<sup>10</sup>** |
| --- | --- | --- | --- |
| **Inclusion Criteria** | **Adults** with nephrotic-range proteinuria ($\geq 500$ mg/d on a 24h urine sample or urinary protein-to-creatinine (UP:C) ratio $\geq 0.5$ g/g on spot urine) were eligible for enrolment at the time of a first clinically indicated kidney biopsy.<br>**Children** with nephrotic-range proteinuria | Steroid therapy naïve children 1–18 years of age presenting with edema and proteinuria $\geq 3+$ by dipstick were eligible for enrolment. | Similar to NEPTUNE |
| **Exclusion Criteria** | Life expectancy $< 6$ months, prior solid organ transplantation, kidney manifestations of systemic disease, patients with pre-enrollment history of VTE or participants with a recorded prescription for anticoagulant or antiplatelet medications | | Similar to NEPTUNE |
| **Treatment at Time of Sample Collection** | 66 patients were on steroids, diuretic and /or immunosuppressive at time of blood collection | Plasma obtained at presentation prior to steroid therapy | Samples collected prior to treatment |
| **Sample Collection** | Blood collected into 0.105 M buffered sodium citrate tubes (0.32% final concentration citrate) | Blood was collected into 0.1 M sodium citrate cell preparation tubes containing a cell separator system ($\sim 0.33\%$ final concentration NaCitrate) | Blood was collected into final concentration 0.32% sodium citrate with 1.45 $\mu\text{M}$ corn trypsin inhibitor (Haematologic Technologies Inc., Essex Junction, VT, USA) |
| **Remission Status Definition** | Complete (UPCR $\leq 0.3$) and partial (UPCR $< 3.5$ plus a $\geq 50\%$ UPCR reduction) NEPTUNE remission criteria were utilized | Steroid-sensitive NS (SSNS) was defined as disease remission following 7 weeks of standard-of-care steroid therapy, whereas steroid-resistant NS (SRNS) was defined as failure to achieve complete remission during this timeframe, as determined by resolution of proteinuria by urine dipstick or UPCR. | Similar to NEPTUNE |

### Nephrotic Syndrome Disease Markers

#### Urine protein

creatinine ratio (UPCR) assays were performed at the NEPTUNE central laboratory for the NEPTUNE cohort. UPCR was determined at the enrolling hospital clinical lab for the PNRC patients. For the Columbus cohort, UPCR was determined using spectrophotometric methods at Nationwide Children’s Hospital clinical laboratory. Plasma albumin was quantified using a bromocresol purple assay kit according to the manufacturer’s instructions (BioAssay Systems, Hayward, CA), as previously described.^10, 39^

### Coagulation Parameters

Thrombin generation assays were performed using Technothrombin TGA kits (Technoclone, Vienna, Austria) and TGA RC Low reagent, read on a Spectramax M2 fluorescent plate reader (Molecular Devices, Sunnyvale, California), as previously described.^10, 11, 15^ TGA generates several hemostasis parameters, including endogenous thrombin potential (ETP), that may be indicative of hypercoagulopathy.^10^ We have previously demonstrated that ETP is strongly correlated with NS severity; thus, ETP has been used to quantify NS-hypercoagulopathy in this study.^10, 11, 15^ Plasma AT antigen was measured by ELISA (Human Antithrombin III ELISA, MyBiosource, San Diego, CA), and validated using latex immunoassay (LIATEST AT III, Diagnostica Stago, Parsippany, NJ; Figure S1). Plasma AT activity was measured using a modified amidolytic fluorescent substrate method, as described previously.^10, 15^ This assay was validated against a commercially available chromogenic assay kit (Hyphen BioMed, Neuville-sur-Oise, France) using standards created by differential mixing of commercially available AT immunodepleted (Affinity Biologicals, Ontario, Canada) and pooled normal (George King Bio-Medical, Overland Park, KS, USA) plasmas (**Figure S1**) and against standards created by addition of an AT neutralizing antibody (Affinity Biologicals, Ontario, Canada).^10^ AT activity and antigen levels were then determined for 8 healthy volunteers and a pooled normal plasma (George King). AT antigen and activity from the NS cohort plasmas were then reported as a percentage of the mean levels of these control plasmas. Human plasma-derived AT (Kybernin® P) for spiking experiments was a kind gift from CSL Behring (Marburg, Germany).

### Meta-Analysis

PubMed and Embase literature searches were used to identify studies published in the English language from 1973 to April 2020. PubMed was searched by Medical Subject Heading terms using the following query: “((((((((nephrotic syndrome) OR minimal change disease) OR focal segmental glomerulosclerosis) OR membranous nephropathy) OR membranous glomerulonephritis) OR membranous glomerulopathy) OR nephrosis)) AND antithrombin.” Embase was searched using the following query: “((((((nephrotic AND (‘syndrome’/exp OR syndrome) OR minimal) AND (‘change’/exp OR change) AND (‘disease’/exp OR disease) OR focal) AND segmental AND (‘glomerulosclerosis’/exp OR glomerulosclerosis) OR membranous) AND (‘nephropathy’/exp OR nephropathy) OR membranous) AND (‘glomerulonephritis’/exp OR glomerulonephritis) OR membranous) AND (‘glomerulopathy’/exp OR glomerulopathy) OR ‘nephrosis’/exp OR nephrosis) AND (‘antithrombin’/exp OR antithrombin) AND (‘embase’/exp OR embase) NOT (‘medline’/exp OR medline).” Bibliographies of included publications were scanned to identify any additional relevant publications. Two authors (JRS and BAK) independently screened the article titles and abstracts to identify studies that reported AT antigen and/or activity values for NS patients. Studies were included if they reported raw data or mean ± variance data. Included studies were also required to report appropriate assay substrate methodologies (i.e. serum was excluded because it is not appropriate for antithrombin assays). If data were not presented as proportional to a reference standard, the data were mathematically converted to a proportion of the healthy control group mean. Publications were excluded if they were reviews, meta-analyses, duplicates, case reports, studies with incomplete data, or if assay methodology was unclear (**Figure S2**). Three hundred sixteen manuscripts were identified in the initial literature search, 110 were fully reviewed, and 27 ultimately included. The included manuscripts’ characteristics are summarized in **Table S1**. Some studies included only adult or pediatric patients, others reported on both. Wherever possible, adult and pediatric patients were analyzed separately. If there was no clear age discrimination in a study including both adults and children, the study was placed in the most appropriate category for the mean age of the study population. AT antigen and activity data generated from the NEPTUNE, PNRC, and Columbus cohort patients with active NS (defined as UPCR >0.3 at the time of plasma collection) were included in the pooled analyses.^10^

### Statistical Analyses

Correlation statistics were performed using Spearman’s Rho correlation coefficient and displayed using scatter plots. Other analyses were performed using GraphPad Prism, version 9.0 (GraphPad Software, San Diego, California). *P*-values <0.05 were considered significant. Meta-analyses were performed using the “meta” R package. Higgins & Thompson’s *I*^*2*^ statistics revealed a relatively high degree of interstudy heterogeneity; thus, Hartung-Knapp-Sidik-Jonkman random effects meta-analyses were used to estimate the pooled difference of mean AT levels between NS patients and controls. Knapp-Hartung adjustment was used to calculate the 95% confidence interval (CI) for the estimated pooled mean difference. A difference of -30% or more between NS patients and controls corresponded to <70% clinical significance threshold.^17, 30^ Forest plots were created to visualize mean differences of individual studies and the pooled mean difference of AT levels. The forest plot x-axes were oriented around 100% (1 IU/mL) for ease of interpretation (e.g. a mean difference between the patients and controls of 0% was plotted as 100%, indicating normal plasma AT).

## RESULTS

### Nephrotic Syndrome Patients

Key NS patient demographic features in the NEPTUNE, PNRC, and Columbus cohorts are summarized in **Table 2**, additional details were previously published.^10, 11^ AT deficiency was observed in all 3 cohorts such that AT antigen and/or activity <70% (<0.7 IU/mL) was observed in 71%, 95%, and 52% of the NEPTUNE, PNRC, and Columbus patients, respectively. AT relationships with plasma albumin, UPCR, and ETP are shown for the NEPTUNE, PNRC, and Columbus cohorts in **Figures 1, 2, and S3**, respectively.

**Table 2:**
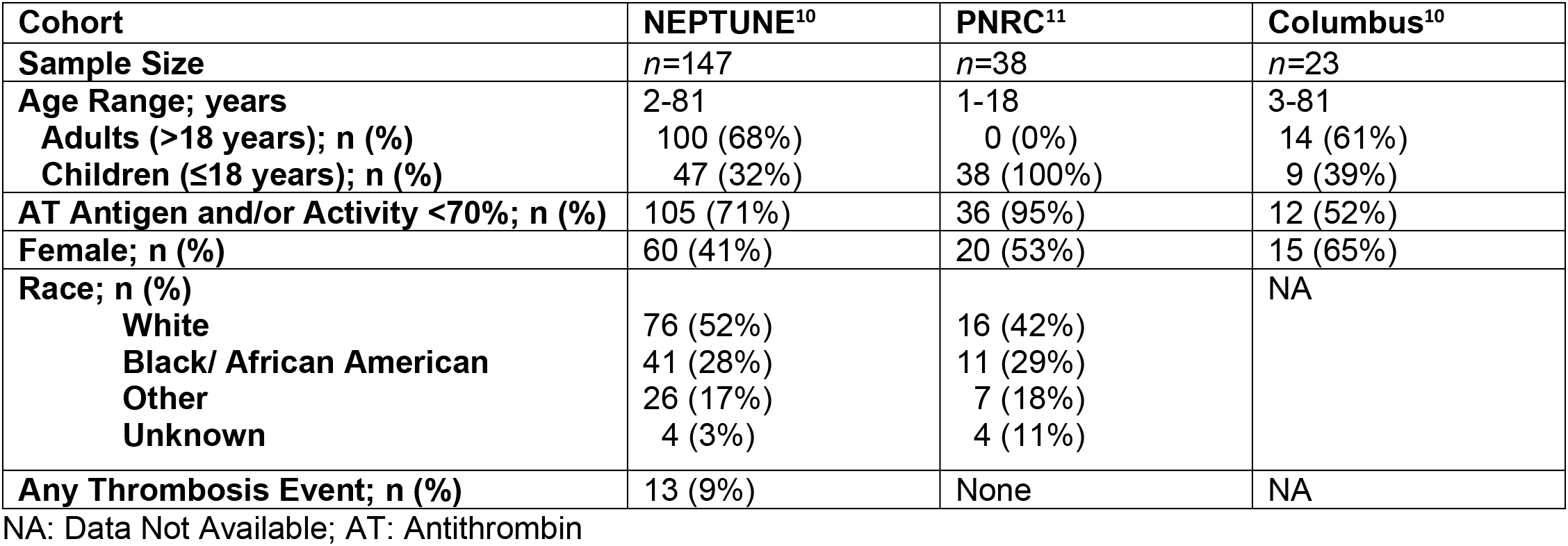
Key patient demographics of the NEPTUNE, Columbus, and PNRC Cohorts.

**Figure 1:**
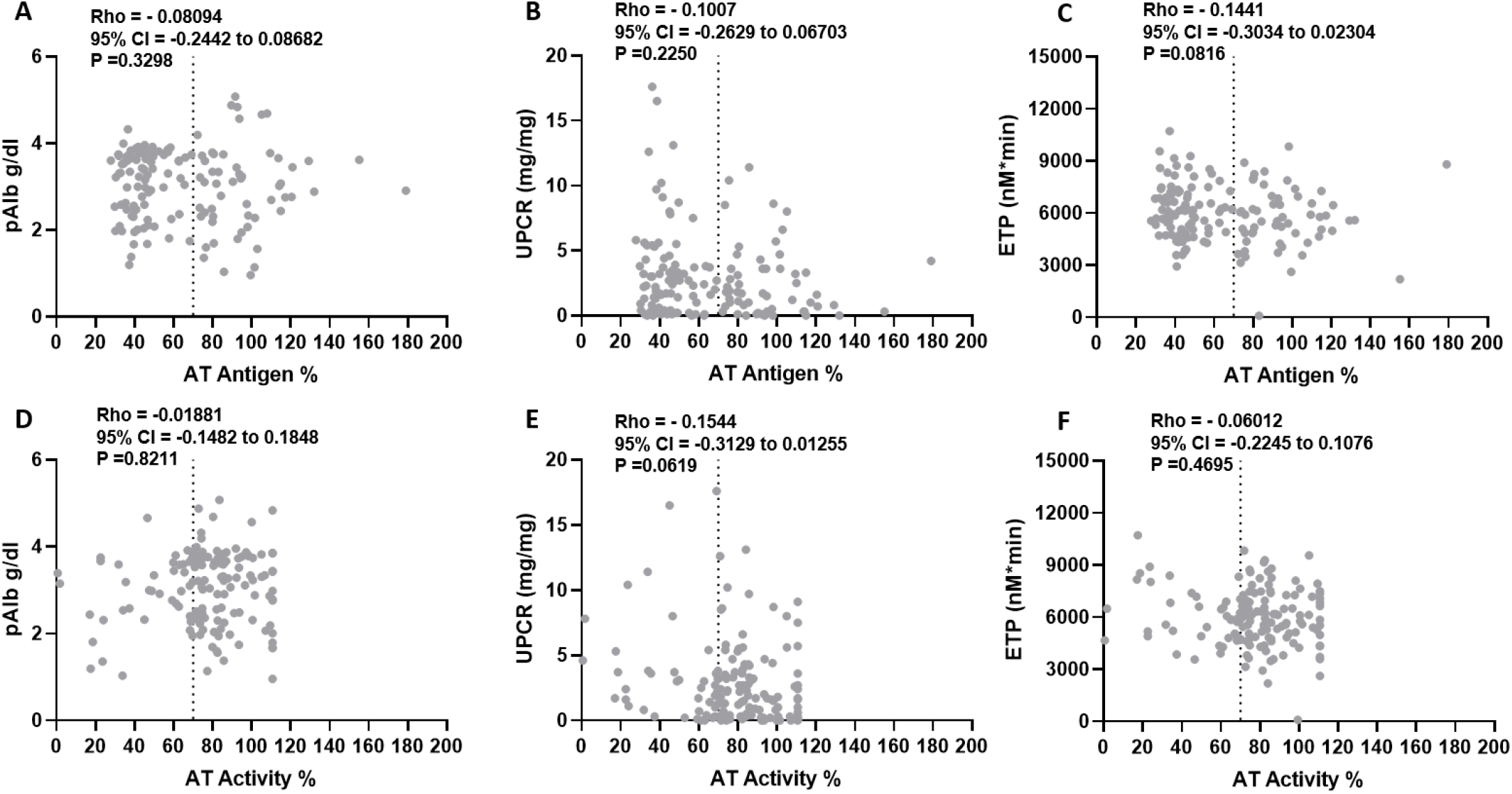
Antithrombin relationships in the NEPTUNE cohort. There is no significant correlation between antithrombin (AT) antigen or activity and plasma albumin (pAlb; **A, D**), urinary protein-to-creatinine ratio (UPCR; **B, E**), or endogenous thrombin potential (ETP; **C, F**) in the NEPTUNE cohort (*n*=147). The vertical dashed line in each panel represents 70% plasma AT, a commonly used threshold to define clinically relevant AT deficiency.

**Figure 2:**
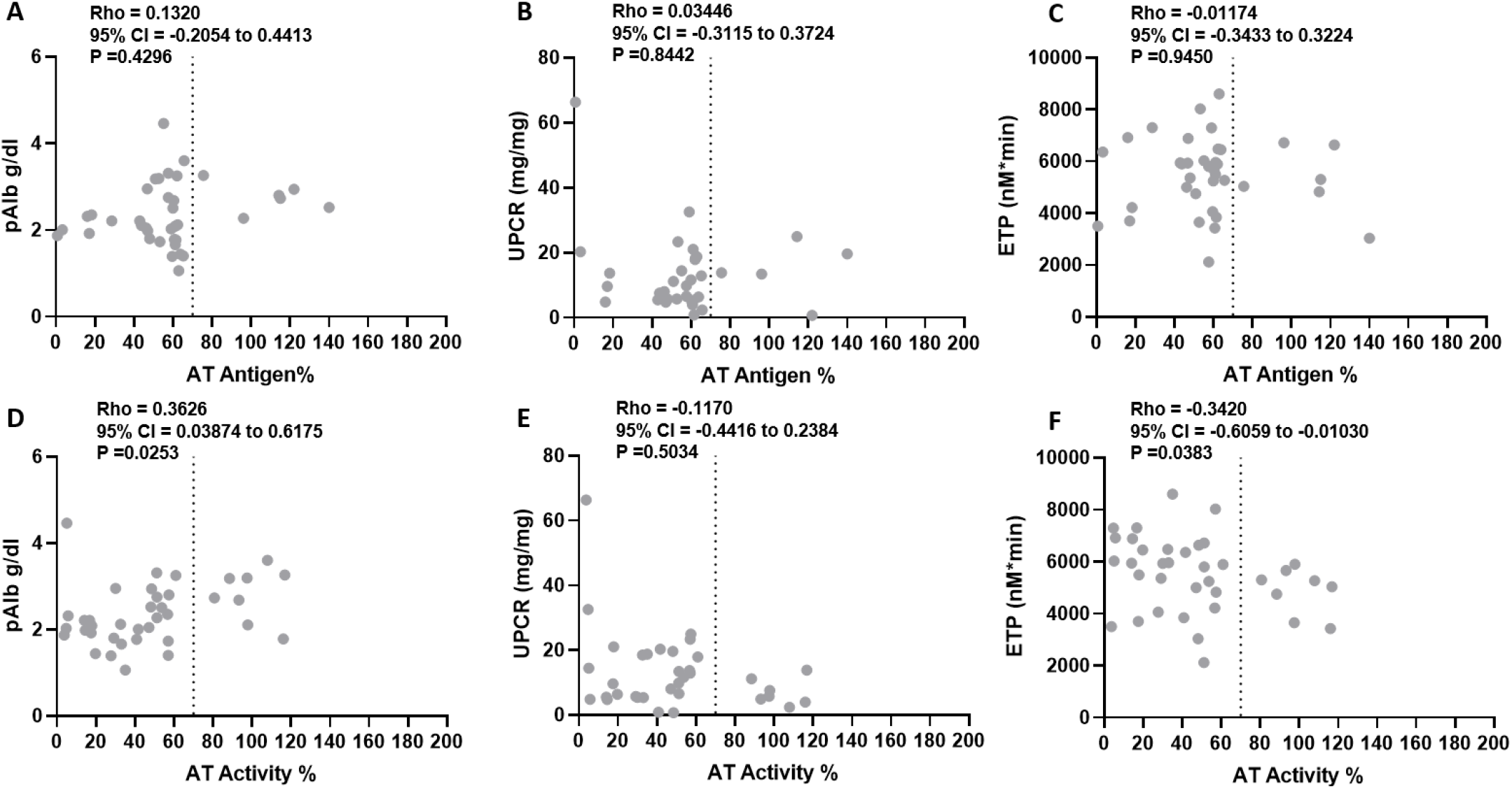
Antithrombin relationships in the PNRC cohort. There is no significant correlation between antithrombin (AT) antigen and plasma albumin (pAlb; **A**), urinary protein-to-creatinine ratio (UPCR; **B**), or endogenous thrombin potential (ETP; **C**) in the PNRC cohort (*n*=38). AT activity is significantly correlated with pAlb (**D**) and ETP (**F**) but not with UPCR (**E**). The vertical dashed line in each panel represents 70% plasma AT, a commonly used threshold to define clinically relevant AT deficiency.

### Proteinuria and Plasma Albumin are not Correlated with Antithrombin Levels

With few exceptions, plasma albumin was not correlated with AT activity or antigen (**Figures 1, 2, and S3)**. AT antigen was correlated with plasma albumin in the Columbus cohort (Rho=0.4201; *P*=0.046; **Figure S3A**) but was not correlated with plasma albumin in any other analysis. Meanwhile, there was a correlation between plasma albumin and AT activity in the PNRC cohort (Rho=0.3626; *P*=0.0253; **Figure 2D**). However, this relationship was not reproducible in the pediatric NEPTUNE subcohort (Rho=-0.1602; *P*=0.2822; **Figure S4D**). Proteinuria (UPCR) was not correlated with AT activity or antigen in any of the cohorts (**Figures 1, 2, S3**). These results suggest that AT levels are not reproducibly correlated with plasma albumin or UPCR during NS.

### Antithrombin Antigen is Correlated with Hypercoagulopathy in Adults with Nephrotic Syndrome

AT levels were not correlated with hypercoagulopathy, as determined by endogenous thrombin potential (ETP), in the NEPTUNE cohort (**Figure 1**). However, the relationship between AT antigen and ETP became significant in the adult NEPTUNE subcohort (Rho=-0.2666; *P*=0.0073; **Figure S5C**) and this relationship was also significant in the smaller, predominantly adult Columbus cohort (Rho=-0.4832; *P*=0.0195; **Figure S3C**). Meanwhile, AT activity was significantly correlated with ETP in the PNRC cohort (Rho=-0.3420; *P*=0.0383; **Figure 2F**). However, as with the plasma albumin data, this relationship was not reproduced in the pediatric NEPTUNE subcohort (Rho=0.0624; *P*=0.677; **Figure S4F**). These data suggest that AT antigen, but not activity, may influence hypercoagulopathy in adult patients. Meanwhile, AT activity is inconsistently correlated with hypercoagulopathy in childhood NS.

### Antithrombin Deficiency does not Explain Nephrotic Syndrome-Hypercoagulopathy

The inconsistent correlation between AT levels and ETP suggested that AT does not directly influence NS-hypercoagulopathy. We thus sought to test the hypothesis that AT deficiency mechanistically drives thrombin generation during NS. AT levels <70% are thought to be a clinically meaningful indicator of VTE risk.^20, 37^ We thus dichotomized our two largest cohorts (NEPTUNE and PNRC) into AT deficient (<70%) vs. non-deficient (≥ 70%) patients to determine if this previously reported cut-off influences NS-hypercoagulopathy. As shown in **Figure 3**, ETP was not significantly influenced by this AT cut-off. Similarly, ETP did not differ by AT quartile in the NEPTUNE cohort (**Figure 3E/F**). Moreover, when representative samples from the most AT deficient NEPTUNE patients (<40% antigen) were spiked to 100% AT *ex vivo*, there was no change in ETP (*P*=0.580; **Figure 3G**). These data strongly suggest that AT deficiency does not significantly contribute to the mechanism underlying NS-hypercoagulopathy.

**Figure 3:**
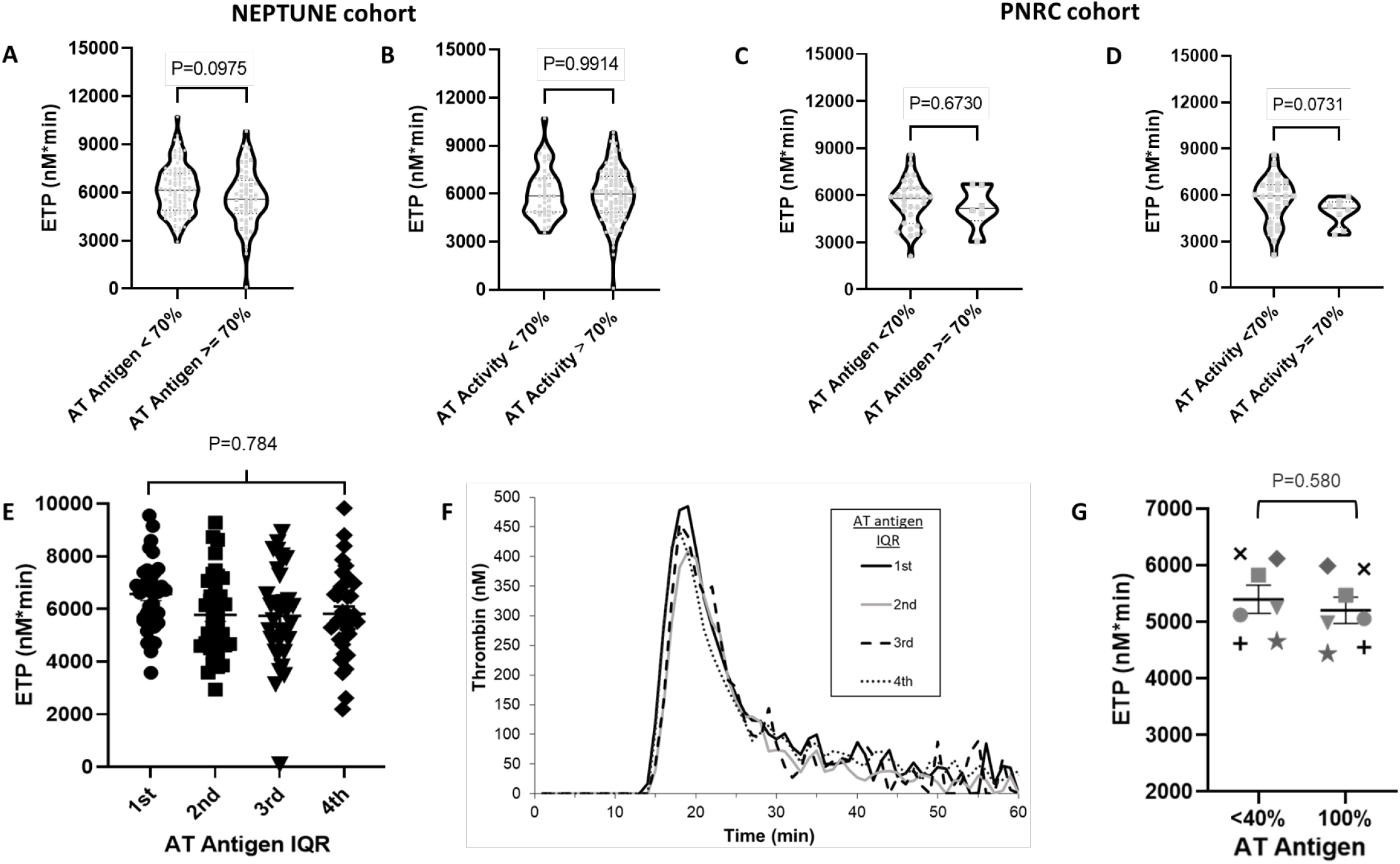
Antithrombin Deficiency does not Explain Nephrotic Syndrome-Hypercoagulopathy. Endogenous thrombin potential (ETP) did not differ between antithrombin (AT) deficient (<70%) and non-deficient (≥ 70%) NEPTUNE (*n*=147; **A, B**) and PNRC (*n*=38; **C, D**) patients. There was also no ETP difference between the NEPTUNE patients when they were distributed into interquartile ranges (IQR) by AT antigen levels (**E, F**). Representative plasma samples from patients in the 1^st^ (lowest) AT antigen quartile (33.81±0.78%) were spiked with human plasma-derived AT to correct AT to 100% did not significantly alter ETP (**G**).

### Antithrombin Deficiency is not a Uniform Feature of Nephrotic Syndrome

To put these new AT data in perspective and evaluate the frequency of NS-related AT deficiency, we next performed meta-analyses. As shown in **Figure 4**, neither AT antigen (Overall Effect: 84.54%; 95% CI: 76.35 to 92.73%) nor activity (96.74%; 95% CI: 78.32 to 115.16%) are expected to routinely fall below 70% in adult NS cases. In contrast, children with NS are more likely to experience AT deficiency, but the overall effect size estimates for both antigen (66.52%; 95% CI: 58.33% to 74.71%) and activity (70.16%; 95% CI: 58.57 to 81.75%) overlap the 70% cut-off. These data indicate that AT deficiency is more likely to be present in children than adults with NS but is not a uniform NS feature in either age group.

**Figure 4:**
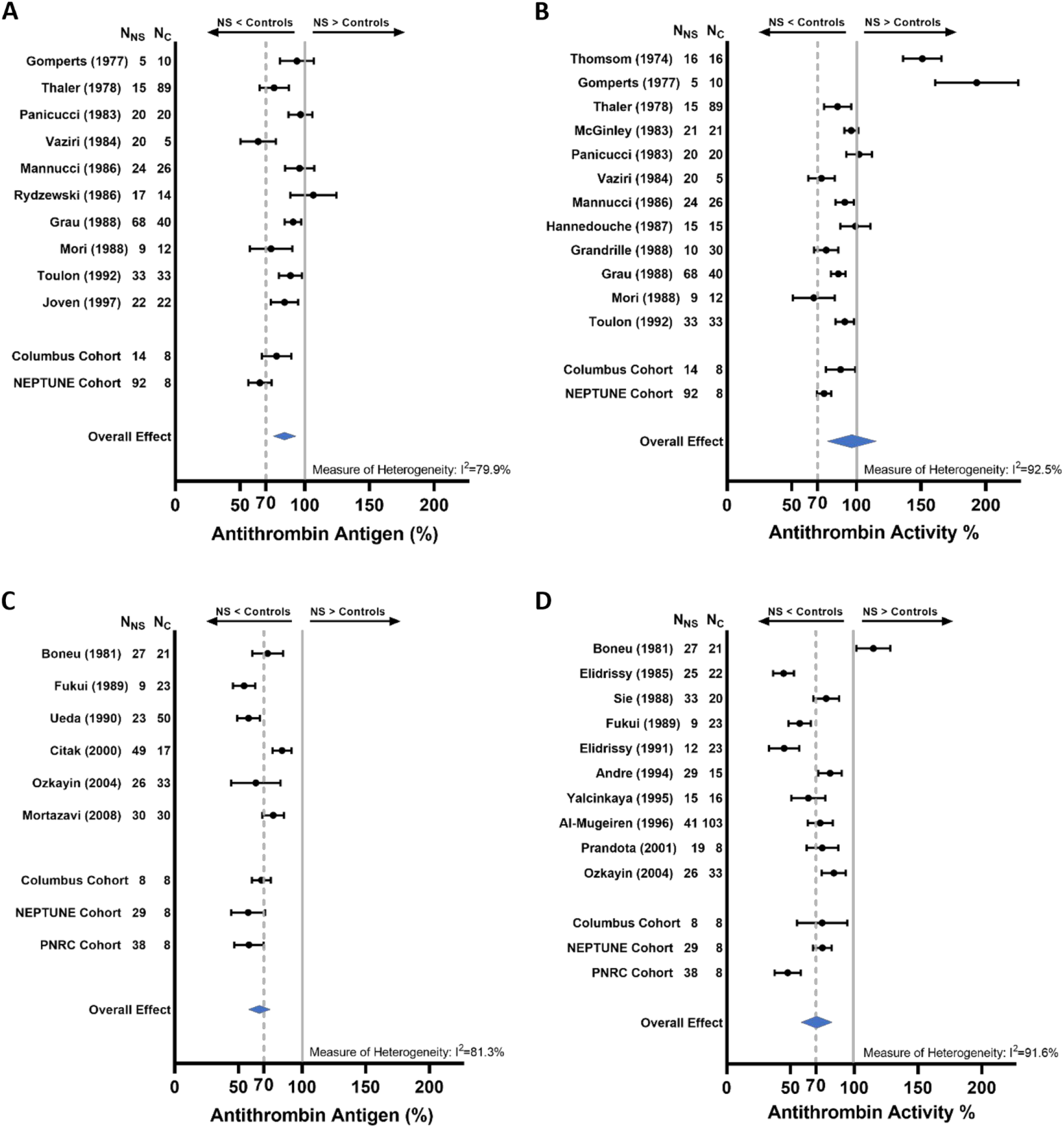
Antithrombin Deficiency is not a Uniform Feature of Nephrotic Syndrome. Based upon the overall effect estimates of this meta-analysis, clinically relevant antithrombin (AT) deficiency (<70%) is not expected in adults with active nephrotic syndrome (NS; **A, B**). AT deficiency is more likely to be observed in children with active NS but the confidence intervals for the overall effect estimates from this meta-analysis overlap with normal AT values (≥ 70%; **C, D**). The vertical dashed line in each panel represents 70% plasma AT, a commonly used threshold to define clinically relevant AT deficiency.

## DISCUSSION

This study evaluated three independent nephrotic syndrome (NS) cohorts to evaluate the likelihood of acquired antithrombin (AT) deficiency in NS and its influence on NS-associated hypercoagulopathy. We found that AT deficiency below a clinically accepted threshold (<70%) was identified in a substantial portion of NS patients. However, AT levels were not strongly associated with either proteinuria or hypoalbuminemia severity. In adult NS, AT antigen levels may be associated with hypercoagulopathy, as determined by thrombin generation. In contrast, the relationship with hypercoagulopathy was an inconsistent finding in childhood NS. However, spiking experiments demonstrated that thrombin generation is not influenced by AT levels. Finally, the meta-analyses demonstrated that acquired AT deficiency is not a uniform feature of NS. Collectively, these data suggest that acquired AT deficiency plays a limited mechanistic role in NS-hypercoagulopathy. Therefore, alternative explanations should be explored to further our mechanistic understanding of hypercoagulopathy and venous thromboembolism (VTE) risk in NS.

VTE is a life-threatening complication of NS and an acquired hypercoagulopathy is thought to be a key driver of NS-associated VTE risk.^1-4, 6, 16-18^ The pathophysiology underlying NS-hypercoagulopathy is likely multifactorial but acquired AT deficiency secondary to urinary AT loss has been postulated to have prominent mechanistic influence.^3, 16-18^ However, there are conflicting data linking NS-associated AT deficiency to both VTE and hypercoagulopathy.^10, 12, 13, 20-26^ This study was thus designed to evaluate the relationships between AT deficiency and both NS disease markers and NS-hypercoagulopathy. The strengths of the study are its inclusion of three independent NS cohorts and its use of meta-analyses incorporating results from these cohorts with the available literature. In all three cohorts, a substantial portion of patients were AT deficient, an observation consistent with numerous prior reports that have been reviewed by our group and others.^2, 3, 6, 16-18^

In contrast to the prevailing hypothesis that AT deficiency is a function of proteinuric loss during NS, we found that AT levels were not correlated with proteinuria severity.^3, 16-18^ Meanwhile, there were weak and inconsistent relationships with plasma albumin levels suggesting that plasma AT levels may be predominantly influenced by compensatory synthesis. However, it is most likely that AT levels result from the combined influence of synthetic compensation and proteinuric loss. Determining which of these mechanisms predominates may require detailed studies designed to simultaneously assess AT synthesis and clearance.^40^

AT antigen, but not activity, levels were correlated with hypercoagulopathy in adult NS patients whereas no consistent relationships were identified in childhood NS. A recent study demonstrated that AT levels were associated with thromboelastography-determined hypercoagulopathy in adult NS.^13^ However, while AT levels were significantly lower in the hypercoagulopathic NS patients, they were not <70% in either group. Meanwhile, we observed no consistent relationship between AT levels and hypercoagulopathy in childhood NS. Moreover, spiking AT back to 100% in representative samples from the most AT deficient patients did not alter ETP. These data suggest that while AT levels (particularly AT antigen in adults) may be a biomarker of NS-hypercoagulopathy, AT deficiency plays only a limited (if any) mechanistic role in NS-hypercoagulopathy. Importantly however, emerging evidence suggest that AT plays an important role in maintaining healthy vascular endothelium via signaling properties that are independent of its coagulation inhibitor function.^41^ It is thus possible that AT deficiency may drive NS-associated VTE risk via vascular mechanisms not captured by TGA or thromboelastography.

While adult patients are more likely to develop VTE than pediatric NS patients, these meta-analyses suggest that AT deficiency is not a uniform feature of adult NS. A general concept in VTE epidemiology is that VTE becomes more common with advancing age.^42^ Moreover, VTE is most common in NS caused by membranous nephropathy, a form of NS rarely observed in children.^2^ Thus, the difference in VTE risk between childhood and adult NS is more likely a combined function of age and NS subtype than of AT deficiency. In contrast, children with NS are more likely to experience AT deficiency but the prediction intervals overlap with the accepted threshold (<70%) defining deficiency. Together these data suggest that AT deficiency is not a consistent NS feature and is thus unlikely to fully explain NS-associated VTE risk.

Limitations of the present studies include inconsistency in the pre-analytic variables used for collection of the plasma samples from the three cohorts, which have been detailed in previous publications.^10, 11^ Importantly, the PNRC samples were collected in gel-containing plasma separator tubes with a sodium citrate content that is slightly higher than the usual coagulation assay standard (∼0.33% vs. 0.32% final concentration, respectively).^11^ It is possible that these conditions led to the observations unique to that dataset (i.e. a correlation between AT activity and ETP). It is also possible that interactions with the gel facilitated conversion to latent AT which is well-known to be inactive.^41, 43, 44^ Alternatively, elastase can irreversibly inactivate AT and elevated elastase levels have been documented in childhood NS.^41, 45, 46^ It is thus possible that these conditions were optimal for pre-analytic elastase-mediated AT inactivation, resulting in a significant correlation not observed in the other pediatric samples. The data utilized in the meta-analyses were collected over nearly a half-century. Although each study used state-of-the-art AT assays for its contemporaneous period, the results from these studies may not be completely comparable since it is likely that AT assay sensitivity and specificity have improved over the past 50 years. This is likely reflected in the relatively high *I*^2^ heterogeneity values, which prompted us to utilized random effects models in the meta-analyses. Moreover, most of the studies compared AT levels to a healthy control group rather than a pooled normal plasma. We thus found it necessary to proportionately estimate AT levels relative to the available standard in each study, which may have led to over- or under-estimation. Despite these limitations there is remarkable consistency in the values across the available studies.

In summary, the studies presented in this manuscript demonstrate that, in contrast to the prevailing hypothesis, AT deficiency plays a limited role in the acquired hypercoagulopathy of NS. Future studies should focus on identifying AT-independent mechanisms that better explain the mechanisms underlying NS-hypercoagulopathy. Meanwhile, the possibility that AT deficiency during NS is associated with a loss of its endothelial cytoprotective functions should be explored as an alternative explanation for the increased risk of VTE during NS.

## Supporting information

Supplemental Tables, Figures, References

PRISMA Checklist

## Data Availability

All data produced in the present study are available upon reasonable request to the authors

## DISCLOSURES

The authors declare no competing interests.

## FUNDING

BAK was supported by grants U54DK083912-05S1, U54DK083912-07S1, K08DK103982, R03DK118315, and R01DK124549 from the National Institute of Diabetes and Digestive and Kidney Diseases (NIDDK) of the National Institutes of Health (NIH). The Nephrotic Syndrome Study Network (NEPTUNE) is part of the Rare Diseases Clinical Research Network (RDCRN), which is funded by the NIH and led by the National Center for Advancing Translational Sciences (NCATS) through its Division of Rare Diseases Research Innovation (DRDRI). NEPTUNE is funded under grant number U54DK083912 as a collaboration between NCATS and the NIDDK. Additional funding and/or programmatic support is provided by the University of Michigan, NephCure Kidney International and the Halpin Foundation. RDCRN consortia are supported by the RDCRN Data Management and Coordinating Center (DMCC), funded by NCATS and the National Institute of Neurological Disorders and Stroke (NINDS) under U2CTR002818.

## ACKNOWLEDGEMENTS

We are indebted to the patient volunteers for their participation in the NEPTUNE, PNRC, and Columbus Cohorts as well as the investigators and staff at each of the enrollment centers who made this study possible. Portions of this work have been presented as abstracts at annual meetings of the American Society of Hematology, International Society on Thrombosis and Haemostasis, and American Society of Nephrology.

## SUPPLEMENTAL MATERIAL

Table of Contents:

**Table S1**

**Figures S1-S5**

## Author contributions

EA performed the meta-analyses and wrote the paper. APW performed experiments and wrote the paper. KJW performed laboratory assays. JRS performed statistics and meta-analyses and wrote the paper. SVP, BHR, and WES enrolled patients and collected samples. The PNRC and NEPTUNE Investigators enrolled patients and collected samples. BAK conceived the study, enrolled patients, collected samples, performed meta-analyses, and wrote the paper. BAK provided financial support. All named authors refined the final draft.

## SUPPLEMENTAL FIGURE LEGENDS

**Figure S1: Antithrombin Assay Validation**. Antithrombin (AT) antigen quantification by latex immunoassay (LIA) and enzyme-linked immunosorbent assay (ELISA) are significantly correlated (*n*=50 NEPTUNE samples; **A**). Similarly, AT activity quantified using amidolytic assays with either chromogenic or fluorogenic reporters provide similar results (**B**). Two types of standard curve were utilized in **B**: On the x-axis standards were created with proportionate mixing of AT immunodepleted plasma and pooled normal plasma whereas the y-axis standards were generated with a set of healthy control plasmas treated with varying concentrations of AT neutralizing antibody (*n*=3-4 samples per point on the standard curve).

**Figure S2: Flow diagram illustrating exclusion and inclusion criteria of publications considered for the meta-analyses**.

**Figure S3: Antithrombin relationships in the Columbus cohort**. Antithrombin (AT) antigen is significantly correlated with plasma albumin (pAlb; **A**) and endogenous thrombin potential (ETP; **C**) but not with urinary protein-to-creatinine ration (UPCR; **B**) in the Columbus cohort (*n*=23). AT activity was not significantly correlated with pAlb (**D**), UPCR (**E**), or ETP (**F**). The vertical dashed line in each panel represents 70% plasma AT, a commonly used threshold to define clinically relevant AT deficiency.

**Figure S4: Antithrombin is not correlated with hypercoagulopathy in the pediatric NEPTUNE subcohort**. Neither antithrombin (AT) antigen (**A, B, C**) or activity (**D, E, F**) are correlated with plasma albumin (pAlb; **A, D**), urinary protein-to-creatinine ratio (UPCR; **B, E**), or endogenous thrombin potential (ETP; **C, F**) in the pediatric NEPTUNE subcohort (*n*=47). The vertical dashed line in each panel represents 70% plasma AT, a commonly used threshold to define clinically relevant AT deficiency.

**Figure S5: Antithrombin antigen is correlated with hypercoagulopathy in the adult NEPTUNE subcohort**. Antithrombin (AT) antigen is correlated with endogenous thrombin potential (ETP; **C**) in the adult NEPTUNE subcohort (*n*=100). There was no significant correlation between AT activity and ETP (**F**) or between either AT antigen or activity with plasma albumin (pAlb; **A, D**) or urinary protein-to-creatinine ratio (UPCR; **B, E**). The vertical dashed line in each panel represents 70% plasma AT, a commonly used threshold to define clinically relevant AT deficiency.

## ADDENDUM 1: PNRC ENROLLING CENTERS

*Nationwide Children’s Hospital and The Ohio State University, Columbus, OH*: J Mahan, H Patel, RF Ransom

*Medical College of Wisconsin, Milwaukee, WI*: C Pan

*The Hospital for Sick Children, Toronto, ON*: DF Geary

*West Virginia University, Charleston, WV*: ML Chang

*University of North Carolina, Chapel Hill, NC*: KL Gibson

*Louisiana State University, New Orleans, LA*: FM Iorember

*University of Iowa Stead Family Children’s Hospital, Iowa City, IA*: PD Brophy

*Children’s Mercy Hospital, Kansas City, MO*: T Srivastava

*Emory University School of Medicine, Atlanta, GA*: LA Greenbaum

## ADDENDUM 2: NEPTUNE ENROLLING CENTERS

*Cleveland Clinic, Cleveland, OH*: K Dell^*^, J Sedor^**^, M Schachere^#^, J Negrey^#^

*Children’s Hospital, Los Angeles, CA*: K Lemley^*^, J Scott^#^

*Children’s Mercy Hospital, Kansas City, MO*: T Srivastava^*^, S Morrison^#^

*Cohen Children’s Hospital, New Hyde Park, NY:* C Sethna^*^, M Pfaiff ^#^

*Columbia University, New York, NY:* P Canetta^*^, A Pradhan^#^

*Emory University, Atlanta, GA:* L Greenbaum^*^, C Wang**, E Yun^#^

*Harbor-University of California Los Angeles Medical Center:* S Adler^*^, J LaPage^#^

*John H. Stroger Jr. Hospital of Cook County, Chicago, IL:* A Athavale^*^, M Itteera

*Johns Hopkins Medicine, Baltimore, MD:* M Atkinson^*^, T Dell^#^

*Mayo Clinic, Rochester, MN:* F Fervenza^*^, M Hogan**, J Lieske^*^, G Hill^#^

*Montefiore Medical Center, Bronx, NY:* F Kaskel^*^, M Ross^*^, P Flynn^#^

*NIDDK Intramural, Bethesda MD:* J Kopp^*^

*New York University Medical Center, New York, NY:* L Malaga-Dieguez^*^, O Zhdanova**, F Modersitzki^#^, L Pehrson^#^

*Stanford University, Stanford, CA:* R Lafayette^*^, B Yeung^#^

*Temple University, Philadelphia, PA:* I Lee^*^, S Quinn-Boyle^#^

*University Health Network Toronto:* H Reich *, M Hladunewich**, P Ling^#^, M Romano^#^

*University of Miami, Miami, FL:* A Fornoni^*^, C Bidot^#^

*University of Michigan, Ann Arbor, MI:* M Kretzler^*^, D Gipson*, A Williams^#^, C Klida^#^

*University of North Carolina, Chapel Hill, NC:* V Derebail^*^, K Gibson^*^, A Froment^#^, F Ochoa-Toro^#^

*University of Pennsylvania, Philadelphia, PA:* L Holzman^*^, K Meyers**, K Kallem^#^, A Swenson^#^

*University of Texas Southwestern, Dallas, TX:* K Sambandam^*^, K Aleman^#^, M Rogers^#^

*University of Washington, Seattle, WA:* A Jefferson^*^, S Hingorani**, K Tuttle**^§^, L Manahan ^#^, E Pao^#^, A Cooper^#§^

*Wake Forest University Baptist Health, Winston-Salem, NC:* JJ Lin*, Stefanie Baker^#^

*Data Analysis and Coordinating Center*: M Kretzler*, L Barisoni**, C Gadegbeku**, B Gillespie**, D Gipson**, L Holzman**, L Mariani**, M Sampson**, J Sedor**, J Zee**, G Alter, H Desmond, S Eddy, D Fermin, M Larkina, S Li, S Li, CC Lienczewski, T Mainieri, R Scherr, A Smith, A Szymanski, A Williams.

*Digital Pathology Committee:* Carmen Avila-Casado (University Health Network, Toronto), Serena Bagnasco (Johns Hopkins University), Joseph Gaut (Washington University in St Louis), Stephen Hewitt (National Cancer Institute), Jeff Hodgin (University of Michigan), Kevin Lemley (Children’s Hospital of Los Angeles), Laura Mariani (University of Michigan), Matthew Palmer (University of Pennsylvania), Avi Rosenberg (Johns Hopkins University), Virginie Royal (University of Montreal), David Thomas (University of Miami), Jarcy Zee (University of Pennsylvania) Co-Chairs: Laura Barisoni (Duke University) and Cynthia Nast (Cedar Sinai).

*Principal Investigator; **Co-investigator^; #^Study Coordinator

^§^Providence Medical Research Center, Spokane, WA

## Notes

### Competing Interest Statement

The authors have declared no competing interest.

### Author Declarations

The IRBs of Nationwide Children's Hospital and the University of Michigan gave ethical approval for this work.

