## Supplemental Tables, Figures, References for "Limited Role of Antithrombin Deficiency in Nephrotic Syndrome-Associated Hypercoagulopathy"

**Table S1: Key elements of studies included in meta-analysis**

| First Author | Publication Year | Study Design | Patients (*N*) | Age Range (Years) | Antithrombin Assay(s) |
|---|---|---|---|---|---|
| **Adult Studies (Majority of Patients >18 years)** | | | | | |
| Thomsom et al[1] | 1974 | CCSC | 16 | 14-65 | Activity |
| Gomperts et al[2] | 1977 | CCSC | 5 | 19-38 | Both Activity and Antigen |
| Thaler et al[3] | 1978 | CCSC | 14 | 9-70 | Both Activity and Antigen |
| McGinley et al[4] | 1983 | CCSC | 21 | 39.4±3.6 | Activity |
| Panicucci et al[5] | 1983 | CCSC | 20 | "Adults" | Both Activity and Antigen |
| Vaziri et al[6] | 1984 | CCSC | 20 | 16-60 | Both Activity and Antigen |
| Mannucci et al[7] | 1986 | CCSC | 24 | 15-70 | Both Activity and Antigen |
| Rydzewski et al[8] | 1986 | CCSC | 17 | 21-61 | Antigen |
| Hannedouche et al[9] | 1987 | CCSC | 15 | 42±18 | Activity |
| Grandrille et al[10] | 1988 | CCSC | 10 | 16-82 | Activity |
| Grau et al[11] | 1988 | CCSC | 68 | 12-72 | Both Activity and Antigen |
| Mori et al [12] | 1988 | CCSC | 9 | 46-76 | Both Activity and Antigen |
| Toulon et al [13] | 1992 | CCSC | 33 | 16-82 | Both Activity and Antigen |
| Joven et al[14] | 1997 | CCSC | 22 | 25-69 | Antigen |
| **Pediatric Studies (Majority of Patients ≤18 years)** | | | | | |
| Boneu et al[15] | 1981 | CCSC | 27 | 3-13 | Both Activity and Antigen |
| Elidrissy et al[16] | 1985 | CCSC | 25 | 3-11 | Activity |
| Sie et al[17] | 1988 | CCSC | 33 | 2-6 | Activity |
| Fukui et al[18] | 1989 | CCSC | 18 | "Children" | Both Activity and Antigen |
| Ueda et al[19] | 1990 | CCSC | 23 | 3-16 | Antigen |
| Elidrissy et al[20] | 1991 | CCSC | 39 | 2-14 | Activity |
| Andre et al[21] | 1994 | CCSC | 29 | 1-14 | Activity |
| Yalcinkaya et al[22] | 1995 | CCSC | 15 | 2.5-13 | Activity |
| Al-Mugeiren[23] | 1996 | CCSC | 41 | 2-14 | Activity |
| Citak et al[24] | 2000 | CCSC | 49 | 1-16 | Antigen |
| Prandota et al[25] | 2001 | PCC | 19 | 1.5-18 | Activity |
| Ozkayin et al[26] | 2004 | CCSC | 26 | 2-15 | Both Activity and Antigen |
| Mortazavi et al[27] | 2008 | CCSC | 30 | 1.4-11 | Antigen |

CCSC: Cross Sectional Case-Control; PCC: Prospective Case-Control

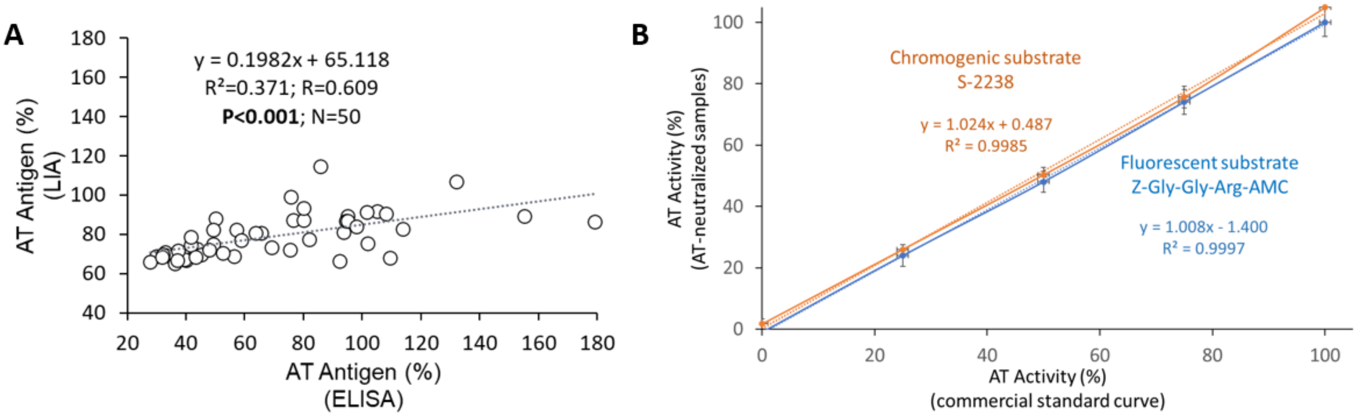


**Figure S1: Antithrombin Assay Validation.** Antithrombin (AT) antigen quantification by latex immunoassay (LIA) and enzyme-linked immunosorbent assay (ELISA) are significantly correlated (*n*=50 NEPTUNE samples; **A**). Similarly, AT activity quantified using amidolytic assays with either chromogenic or fluorogenic reporters provide similar results (**B**). Two types of standard curve were utilized in **B**: On the x-axis standards were created with proportionate mixing of AT immunodepleted plasma and pooled normal plasma whereas the y-axis standards were generated with a set of healthy control plasmas treated with varying concentrations of AT neutralizing antibody (*n*=3-4 samples per point on the standard curve).

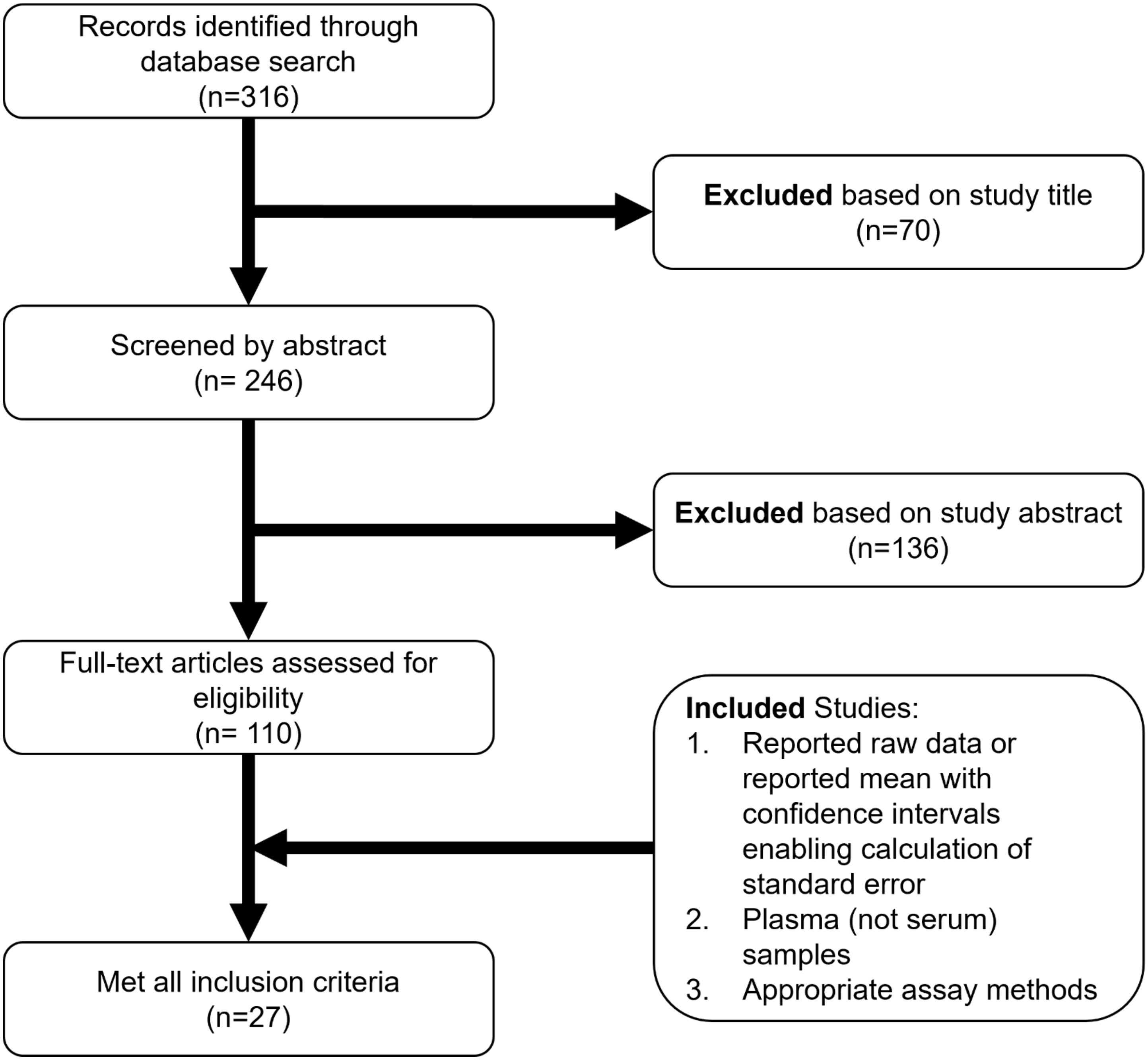


**Figure S2: Flow diagram illustrating exclusion and inclusion criteria of publications considered for the meta-analyses.**

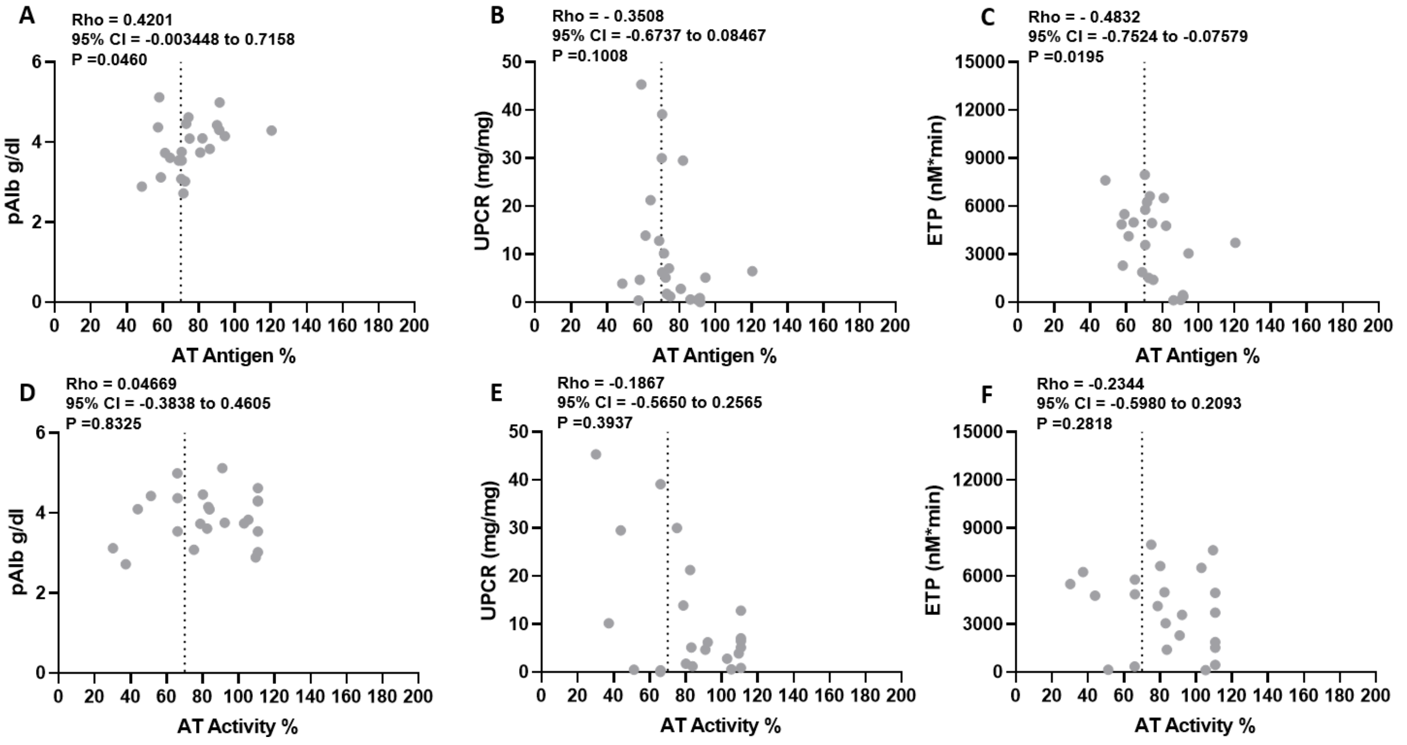


**Figure S3: Antithrombin relationships in the Columbus cohort.** Antithrombin (AT) antigen is significantly correlated with plasma albumin (pAlb; **A**) and endogenous thrombin potential (ETP; **C**) but not with urinary protein-to-creatinine ration (UPCR; **B**) in the Columbus cohort (*n*=23). AT activity was not significantly correlated with pAlb (**D**), UPCR (**E**), or ETP (**F**). The vertical dashed line in each panel represents 70% plasma AT, a commonly used threshold to define clinically relevant AT deficiency.

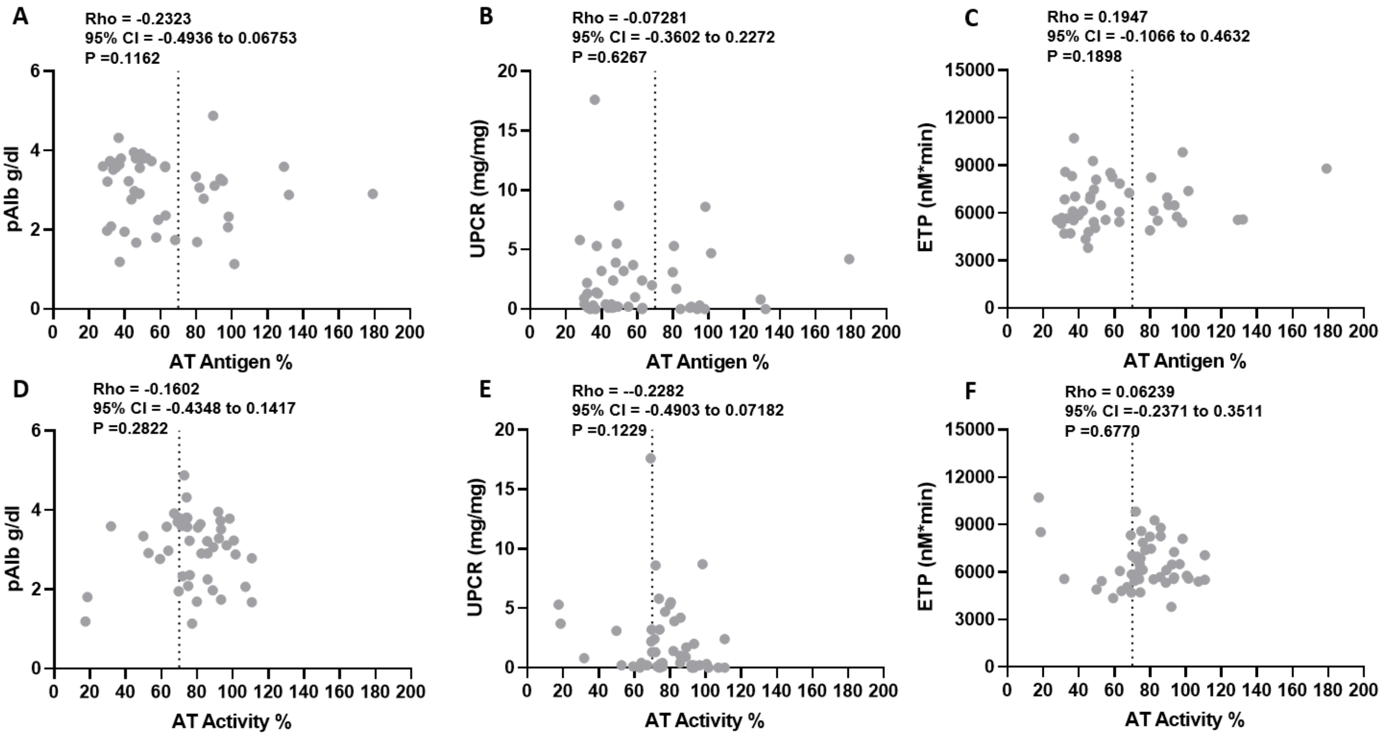


**Figure S4: Antithrombin is not correlated with hypercoagulopathy in the pediatric NEPTUNE subcohort.** Neither antithrombin (AT) antigen (**A**, **B**, **C**) or activity (**D**, **E**, **F**) are correlated with plasma albumin (pAlb; **A**, **D**), urinary protein-to-creatinine ratio (UPCR; **B**, **E**), or endogenous thrombin potential (ETP; **C**, **F**) in the pediatric NEPTUNE subcohort (*n*=47). The vertical dashed line in each panel represents 70% plasma AT, a commonly used threshold to define clinically relevant AT deficiency.

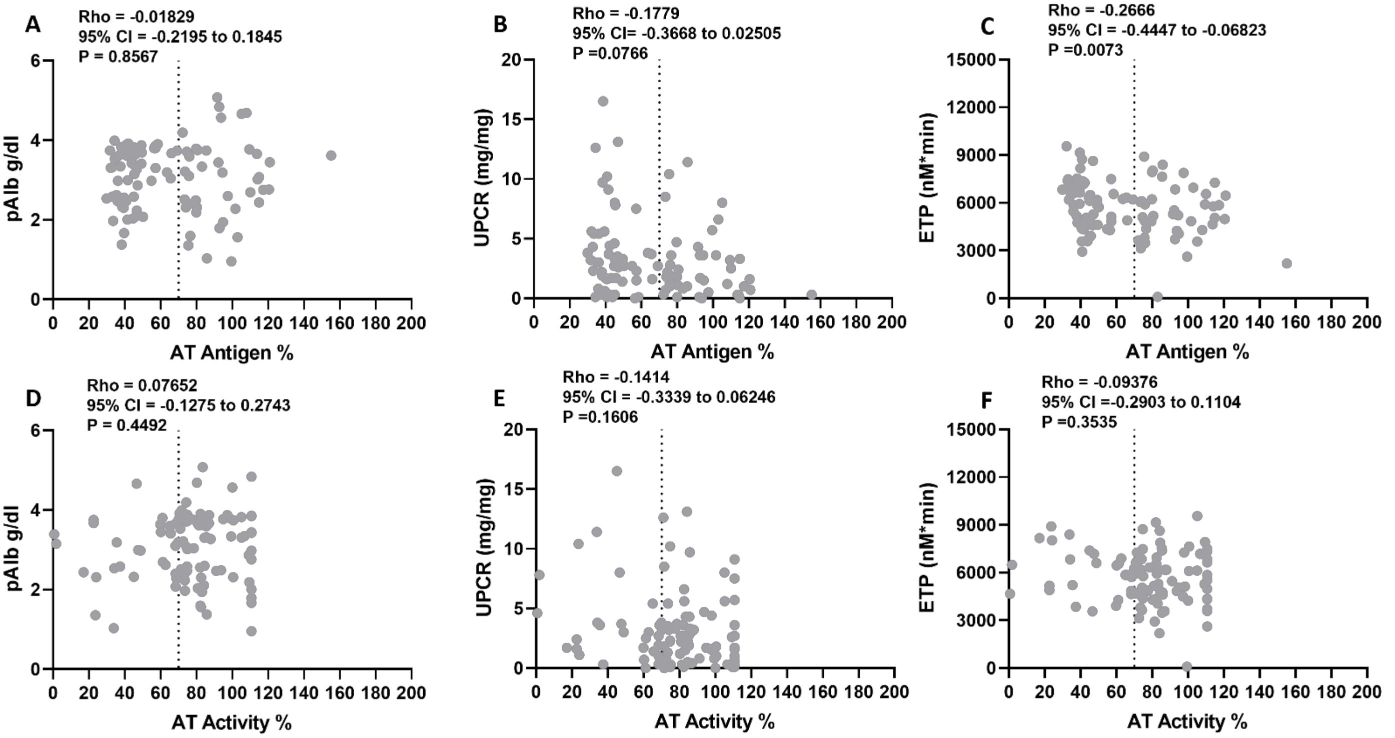


**Figure S5: Antithrombin antigen is correlated with hypercoagulopathy in the adult NEPTUNE subcohort**. Antithrombin (AT) antigen is correlated with endogenous thrombin potential (ETP; **C**) in the adult NEPTUNE subcohort (*n*=100). There was no significant correlation between AT activity and ETP (**F**) or between either AT antigen or activity with plasma albumin (pAlb; **A**, **D**) or urinary protein-to-creatinine ratio (UPCR; **B**, **E**). The vertical dashed line in each panel represents 70% plasma AT, a commonly used threshold to define clinically relevant AT deficiency.
