## Supplementary material for "Limited Role of Antithrombin Deficiency in Nephrotic Syndrome-Associated Hypercoagulopathy": PRISMA Checklist

CJASN
Clinical Journal of the American Society of Nephrology

JASN
JOURNAL OF THE AMERICAN SOCIETY OF NEPHROLOGY

Kidney360

Manuscript ID:

### Adapted* PRISMA Checklist for Systematic Reviews

| Section/Topic | Checklist Item | Response |
|---|---|---|
| **Background** | An explicit statement of questions being addressed with reference to participants, interventions, comparisons, outcomes, and study designs (PICOS). | Yes |
| **METHODS** | | |
| **Protocol and Registration** | Indicates if a review protocol exists and if and when it can be accessed. | NA |
| | Indicates if the protocol has been registered, and if so, provides registration number. | N/A |
| **Eligibility Criteria** | Specifies study characteristics (e.g., PICOS, length of follow-up) and reports characteristics (e.g. years considered, language, publication status used as criteria for eligibility. | Yes |
| **Information Sources** | Describes all information sources (e.g. database with dates of coverage, contact with study authors to identify additional studies) in the search and the date last searched. | Yes |
| **Search** | Presents full electronic search strategy for at least one database, including any limits used, such that it could be repeated. | Yes |
| **Study Selection** | States the process for selecting studies (e.g. screening, eligibility, included in systematic review and/or inclusion in meta-analyses). | Yes |
| **Data Collection Process** | Describes the method of data extraction from reports (e.g., piloted forms, independently, in duplicate) and any processes for obtaining and confirming data from investigators. | Yes |
| **Data Items** | Lists and defines all variables for which data were sought (e.g., PICOS, funding sources) and any assumptions and simplifications made. | Yes |
| **Risk of Bias in Individual Studies** | Describes methods used for assessing risk of bias of individual studies (including specification of whether this was done at the study or outcome level), and this information was used in any data synthesis. | No |
| **Summary Measures** | States the principal summary measures (e.g., risk ratios, difference in means). | Yes |
| **Synthesis of Results** | Describes the methods of handling data and combining results of studies, if done, including measures of consistency ($I^2$) for each meta-analyses. | Yes |
| **RESULTS** | | |
| **Study Selection** | The numbers of studies screened, assessed for eligibility, and included in the review, with reasons for exclusion at each stage | Yes |
| **Study Characteristics** | Characteristics for which data were extracted from each study (e.g., study size, PICOS, follow-up period). | Yes |
| **Risk of Bias Across Studies** | Presents results of any assessment of risk of bias across studies. | No |
| **Additional Analyses** | Gives results of secondary analyses (e.g., sensitivity or sub-group analyses, meta-regression). | No |

**Additional Details:**

*This checklist has been adapted by the ASN journals for author and editor use.

*November 2021*
